# Current State and Predicting Future Scenario of COVID-19 Pandemic for Highly Infected Nations

**DOI:** 10.1101/2020.03.28.20046235

**Authors:** Nandan L. Patil

## Abstract

Since the first report of COVID-19 from Wuhan China, the virus has rapidly spread across the globe now presently reported in 177 countries with positive cases crossing 400 thousand and rising. In the current study, prediction is made for highly infected countries by a simple and novel method using only cumulative positive cases reported. The rate of infection per week (*R*_*w*_) coefficient delineated three phases for the current COVID-19 pandemic. All the countries under study have passed Phase 1 and are currently in Phase 2 except for South Korea which is in Phase 3. Early detection with rapid and large-scale testing helps in controlling the COVID-19 pandemic. Staying in Phase 2 for longer period would lead to increase in COVID-19 positive cases.

## Introduction

Since the first report of COVID-19 in Wuhan province in the month of December, the virus has spread across the globe through foreign travelers at rapid pace. The SARS-CoV2 is transmitted human to human among close contacts (within about 6 feet) [1] and has mean incubation period of 6.4 days with range of 2.1 to 11.1 days [2]. Asymptotic spread has made detection of virus very difficult. Reverse transcription PCR is widely used for detecting COVID- 19 virus which may take few hours and days to obtain results. Observing threat to public health, World Health Organization (WHO) has declared novel coronavirus (2019-nCoV) outbreak to be “public health emergency of international concern” on Jan. 30, 2020 [3] and on the 11th of March the disease was declared a global pandemic [4].

Many prediction models are being proposed for the current pandemic [4][5] namely SIRD model [7], GLEaM [8], Mechanistic-statistical SIR [8], SEIRUS model [9]. These models are often complex and have multiple parameters. Here we propose a simple prediction method for the current COVID-19 pandemic based on only reported positive COID-19 cases. The current study is aimed to predict future scenario of eight highly infected countries namely Italy, Spain, Germany, France, United States, Iran, South Korea and India.

## Methods

The data of confirmed positive COVID-19 cases up to March 27, 2020 was obtained from Wikipedia and Worldmeters [11][12]. Cumulative Positive cases and Total **P**ositive **C**ases for nth **W**eek (*PCW*_*n*_) was calculated. *PCW* is calculated considering a week to be from Saturday to next Friday. A new coefficient Rate of infection per week (***R***_***w***_) was calculated as follows,

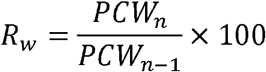

where,

*PCWn* is Total Positive cases for n^th^ week

*PCD*_*n*-1_ is the Total Positive cases for n-1^th^ week

The data was divided into four weeks Feb 22-Feb 28, Feb 29-Mar 6, Mar 7-Mar 13, Mar 14-Mar 20 and Mar 21-Mar 27. All the cases before Feb 22 were grouped into one interval. The epidemic is assumed to follow normal distribution and virus is locally transmitted. All the predictions are made assuming the current control measures.

The factors such as incubation period, time taken to test a sample, time taken to identify primary and secondary contacts have low degree of influence on *R*_*w*_ coefficient and is thus suitable for predicting future scenario of epidemic.

## Results and Discussion

The association of *R*_*w*_ with normal curve is presented in Table 1. When the *R*_*w*_ is above 100, *PCW* is continuously increasing (Figure 1), the peak of normal curve has *R*_*w*_ equal to 100 where there is constant increase in *PCW* and when *R*_*w*_ is below 100, *PCW* starts decreasing.

**Table 1.** Relationship between *R*_*w*_ and *PCW*.

| $R_w$ value | Total Positive Cases per Week ($PCW$) |
| --- | --- |
| > 100 | Increasing |
| 100 | Stable |
| < 100 | Decreasing |

**Figure 1.**
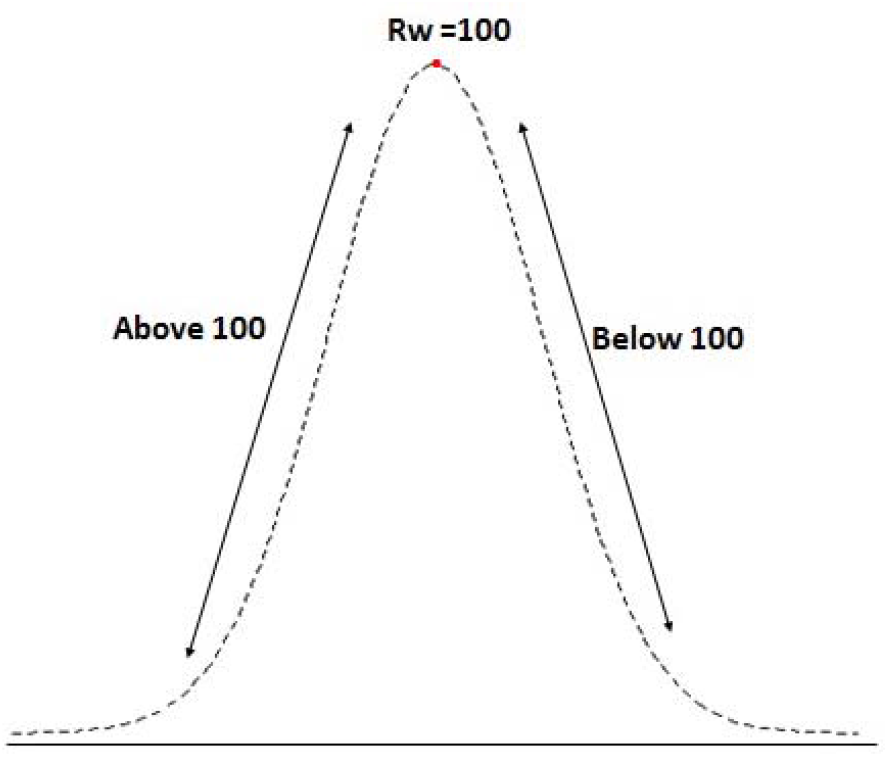
Normal curve defined with respect to *R*_*w*_.

Rw for each country delineates three Phases for the current COVID-19 outbreak caused by SARS CoV 2 (Figure 2, Sup. Table 1).

**Figure 2.**
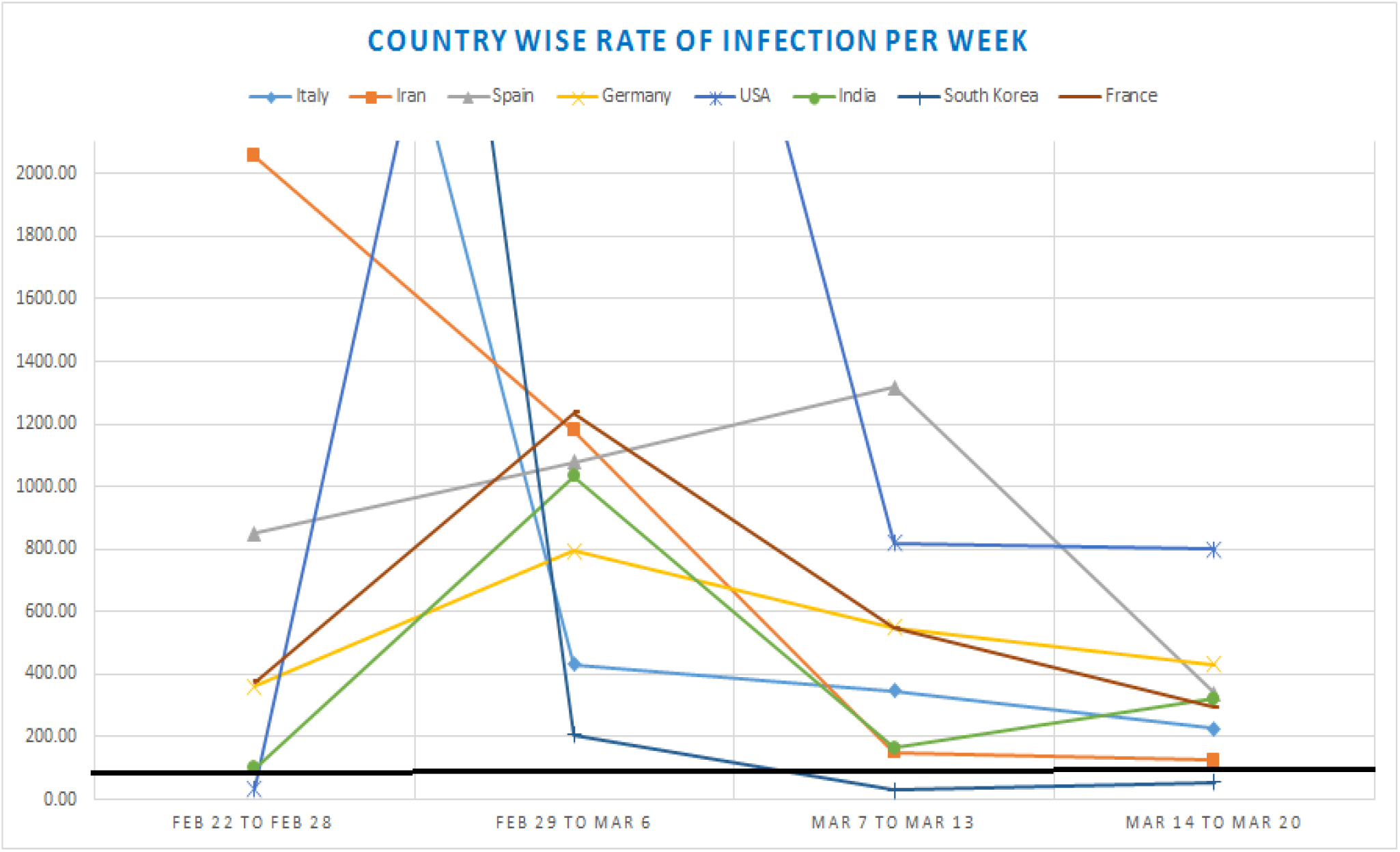
Country-wise Rate of Infection per Week (*R*_*w*_) The bold black line represents Critical Point.

### Phase 1

This phase is defined by sudden spike of which is due to magnitude of positive cases reported. Initially a smaller number of people are tested due to longer incubation period of virus, (2.1 to 11.1 days with mean of 6.4 days) [2] which leads to low positive cases. This is followed by higher testing samples reporting high number of positive cases.

### Phase 2

In this phase the is above 100 but is decreasing at a certain rate. There is currently popular opinion about flattening the epidemic curve to reduce the load on healthcare sector [13][14]. The decay of decides the epidemic curve, a fast decay helps in flattening the curve whereas slow decay would lead to steep normal curve.

### Phase 3

In this phase, is below 100 and approaching zero. Rw equal to 100 is the ‘<u>Critical Point</u>’ as value above this point would increase and value below this point would decrease. Thus, this is a Safe Phase.

### Italy

Italy reported its first case on 31^st^ January and had only 20 cases as of February 21. The Phase 1 was from Feb 22–Feb 28 where is saw a drastic rise in to 869 and recorded of 4345. It is now in Phase 2 and predicted to be currently at the peak of epidemic curve with of 134.45. It is predicted to be in Phase 3 (Safe Phase) in the next week Mar 28-Apr 3 with decreasing and below 100.

**Figure 3.**
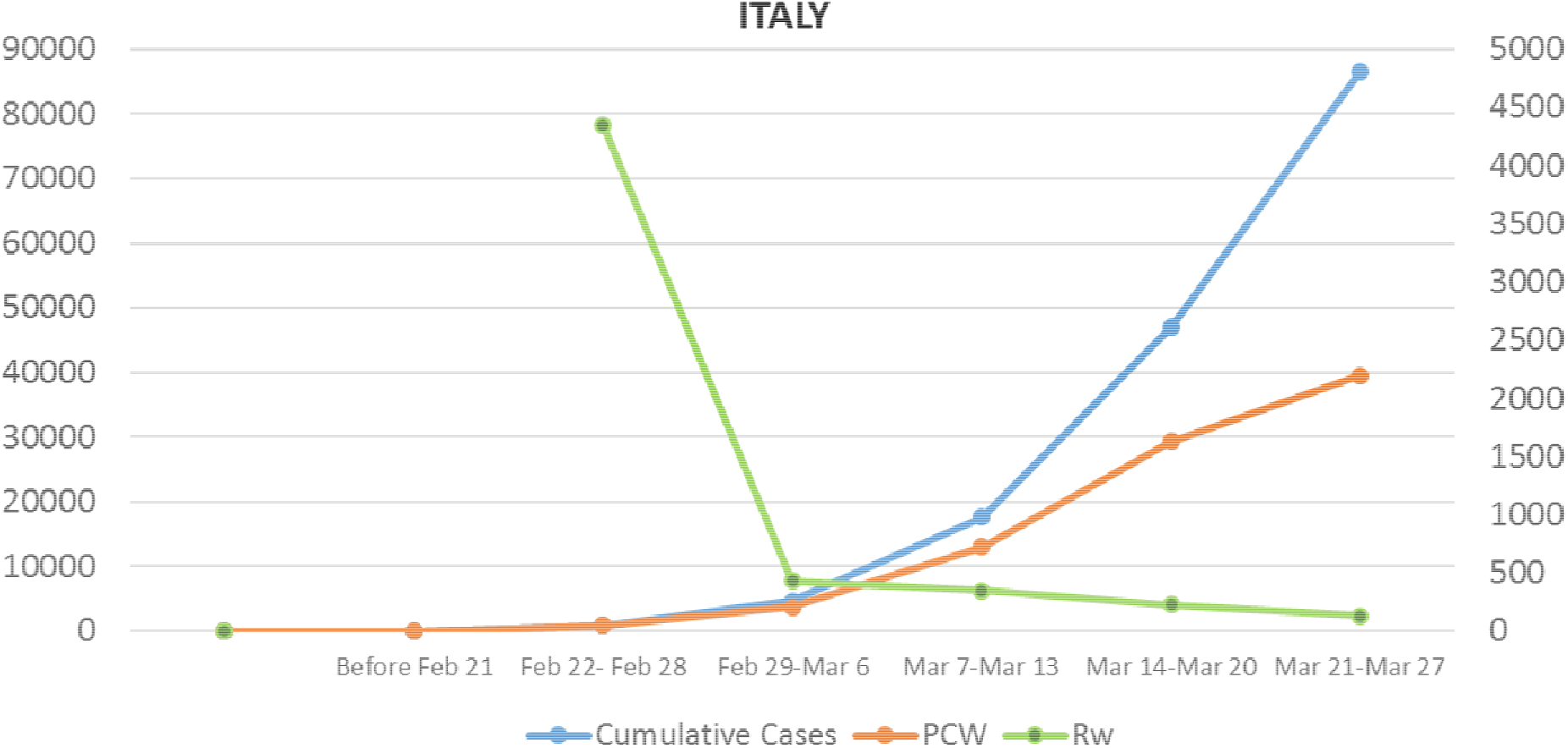
Graphical representation of positive cases for Italy. (The secondary Y axis represents unit for)

### United States

US saw its first case on of COVID-19 on Jan 20, 2020. US observed its Phase 1 from Feb 29 to Mar 6 with of 4660. Now currently it is in Phase 2 with a slow decay of. This slow decay would take form of steep normal curve leading to huge increase in and is expected to reach 400,000 positive cases within Apr 3. Immediate actions need to be undertaken to control the disease in United States. WHO said US could be next epicenter for COVID-19 [15].

**Figure 4.**
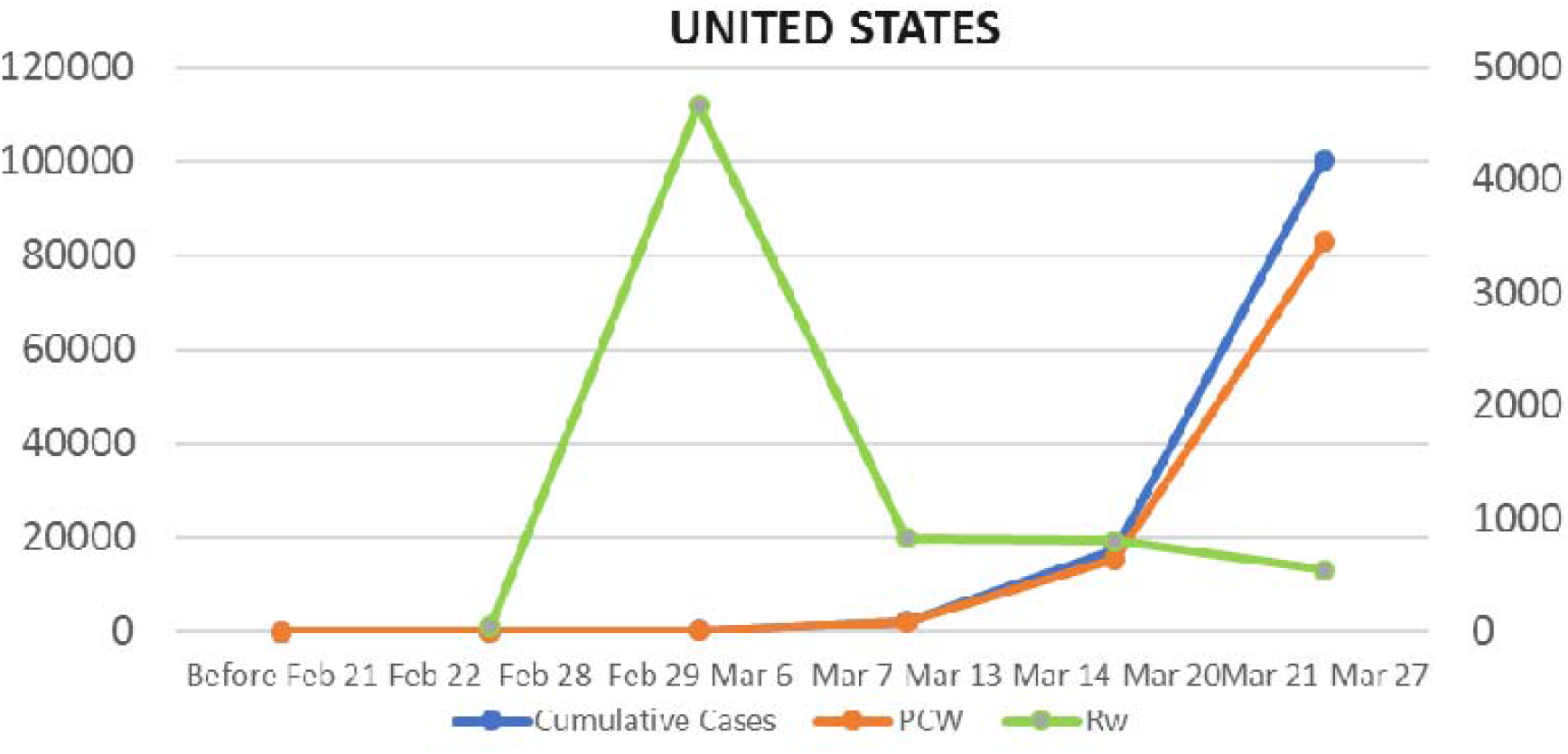
Graphical representation of positive cases for United States. (The secondary Y axis represents unit for)

### Spain

Spain reported its first positive case on 31 January. The Phase 1 was during Mar 7-Mar 13 with of 1316.35. It is predicted to be at the peak of curve in next week and would recorded highest PCW.

**Figure 5.**
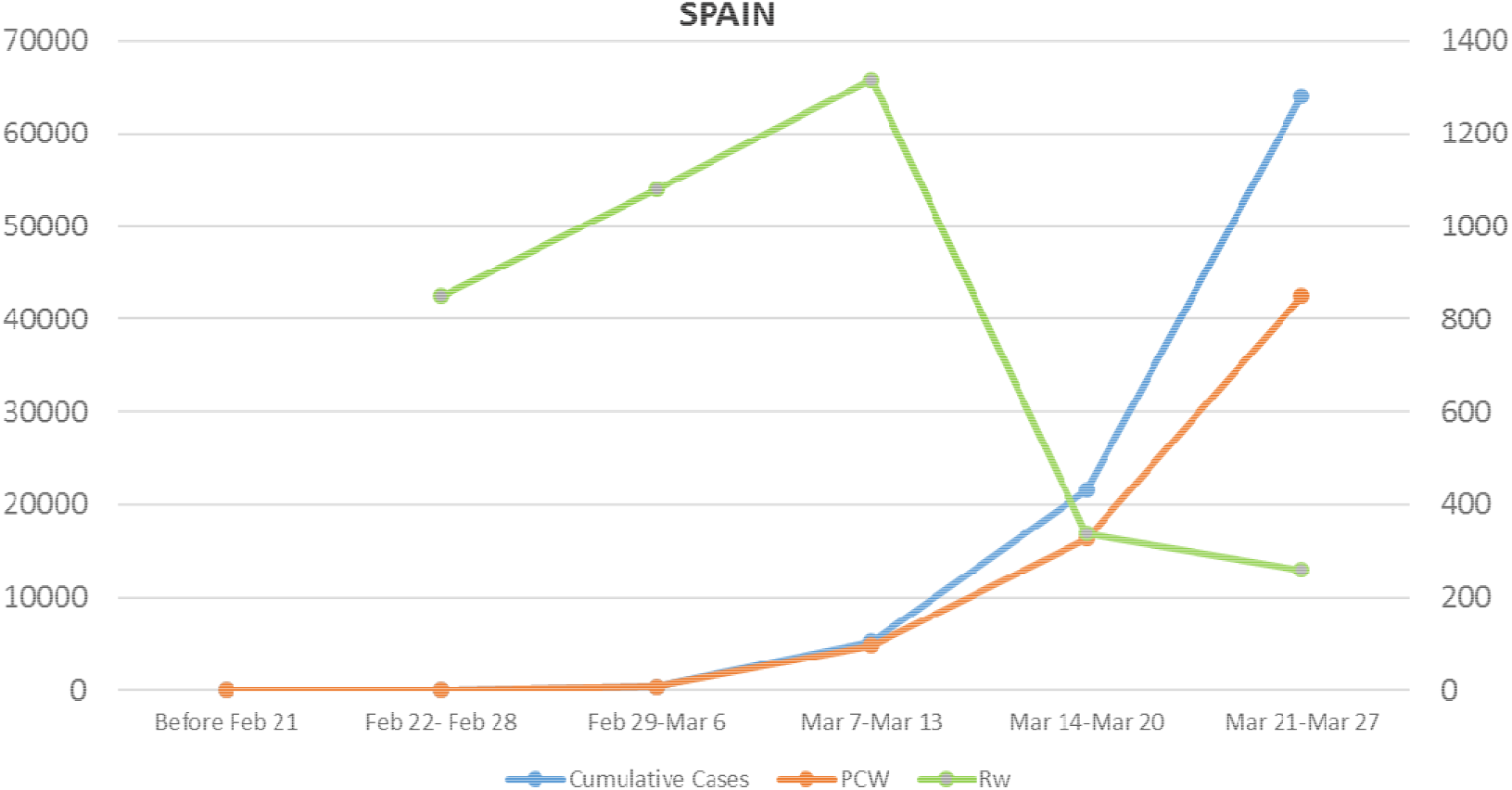
Graphical representation of positive cases for Spain. (The secondary Y axis represents unit for)

### Germany

Germany had its first case on January 27 and passed Phase 1 from Feb 29-Mar 6 with lowest (793.10) compared to other countries under study. It is currently in Phase 2 with a gradual decrease in. Germany is predicted to be at the peak of normal curve with highest for next week (Mar 28-Apr 3) and to be in Safe Phase by 10 April.

**Figure 6.**
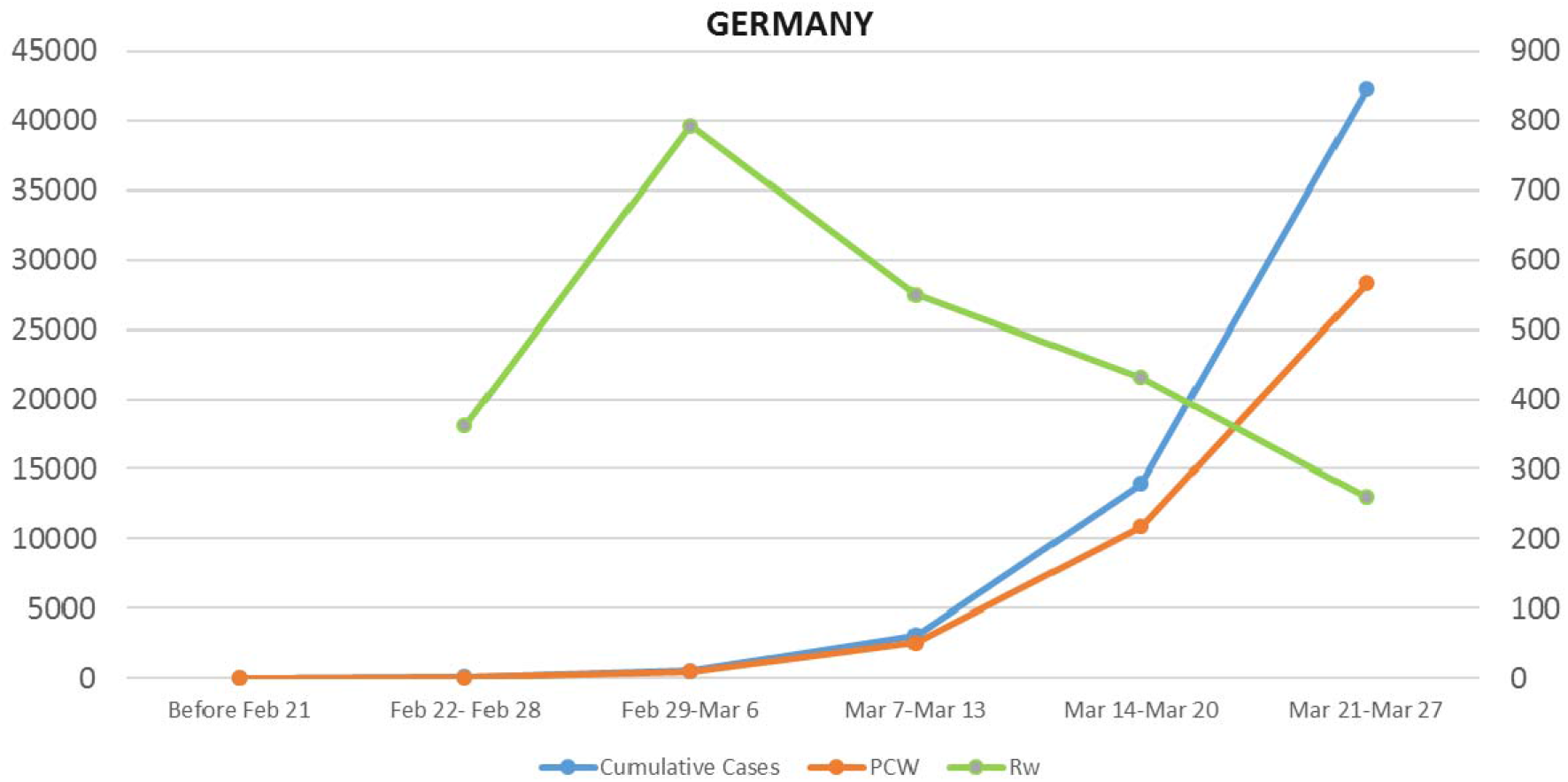
Graphical representation of positive cases for Germany. (The secondary Y axis represents unit for)

### France

France reported its first case on January 24 and passed Phase 1 from Feb 29 to Mar 6. Now it is in Phase 2 and is predicted to have above 100 for next week.

**Figure 7.**
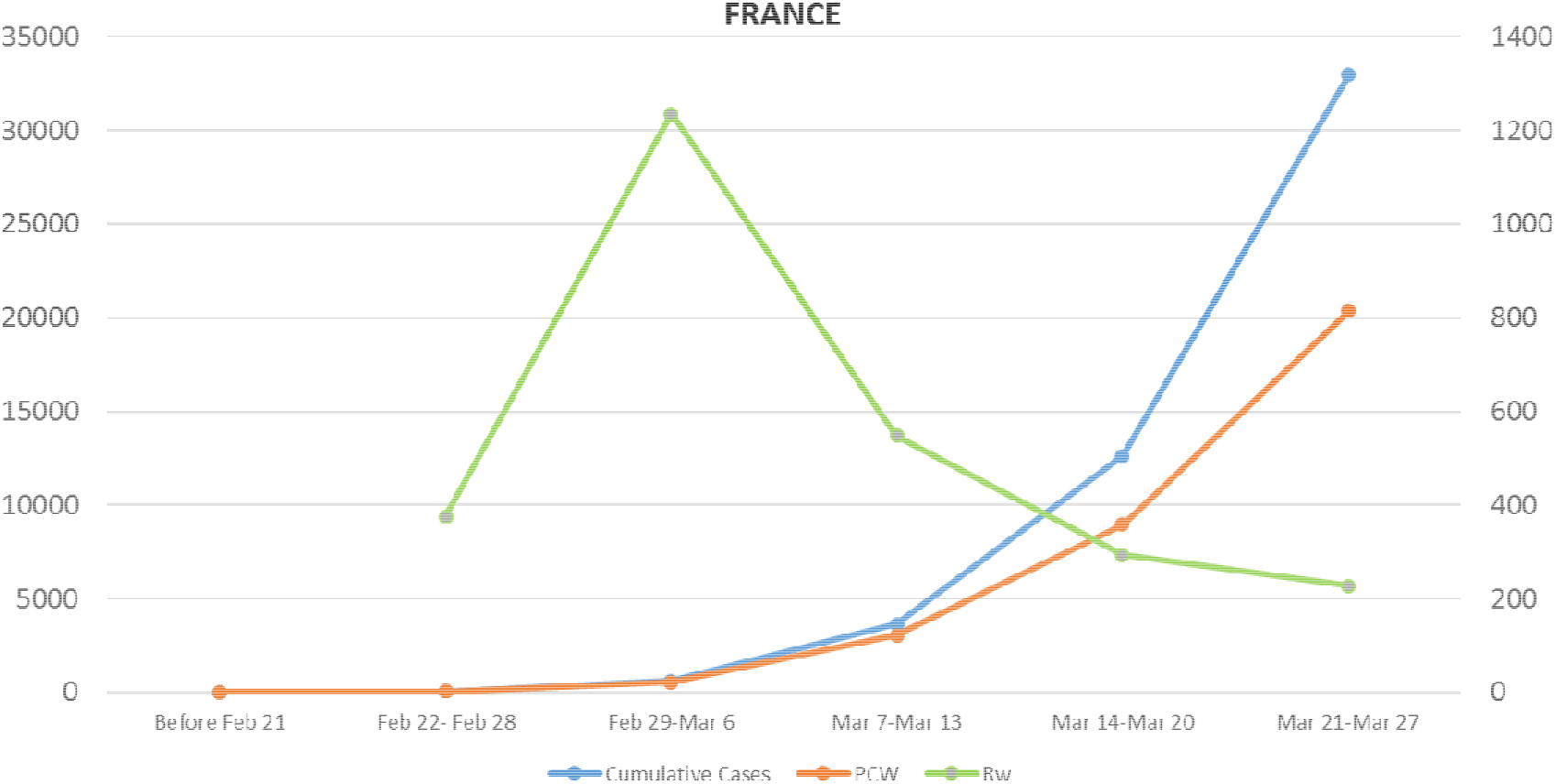
Graphical representation of positive cases for France. (The secondary Y axis represents unit for)

### Iran

Iran reported first positive case on February 19. It has passed Phase 1 during Feb 22-Feb 28 and is currently in Phase 2. It is predicted to have above 100 for the next week.

**Figure 8.**
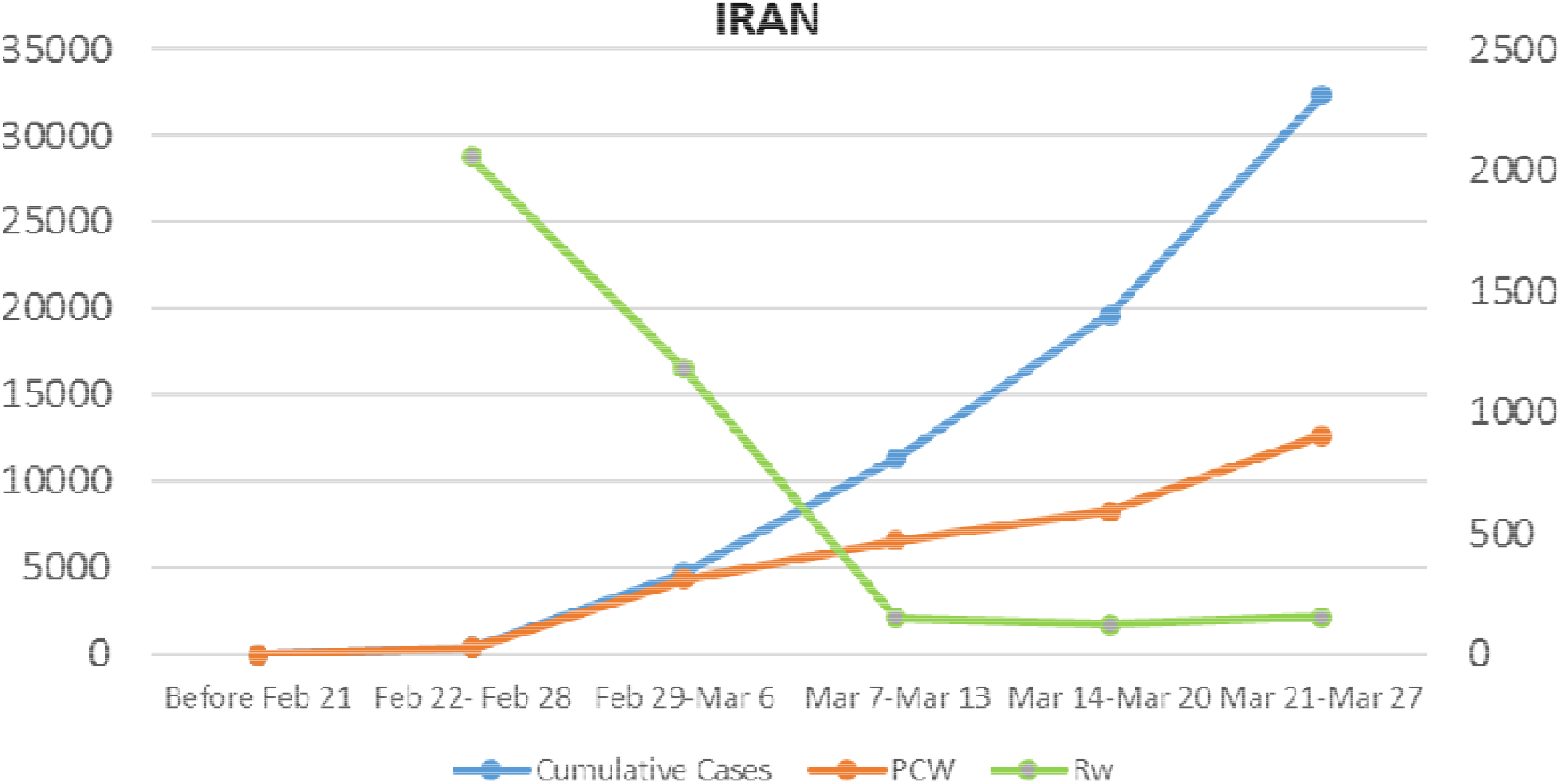
Graphical representation of positive cases for Iran. (The secondary Y axis represents unit for)

### South Korea

South Korea passed Phase 1 on Feb 22-Feb 28 with of 7617.86 highest compared to other countries. This high value can be attributed to large scale testing conducted. It is the only country to pass Phase 2 (Feb 29-Mar 6) and now is in Phase 3 for three weeks with both continuously decreasing.

**Figure 9.**
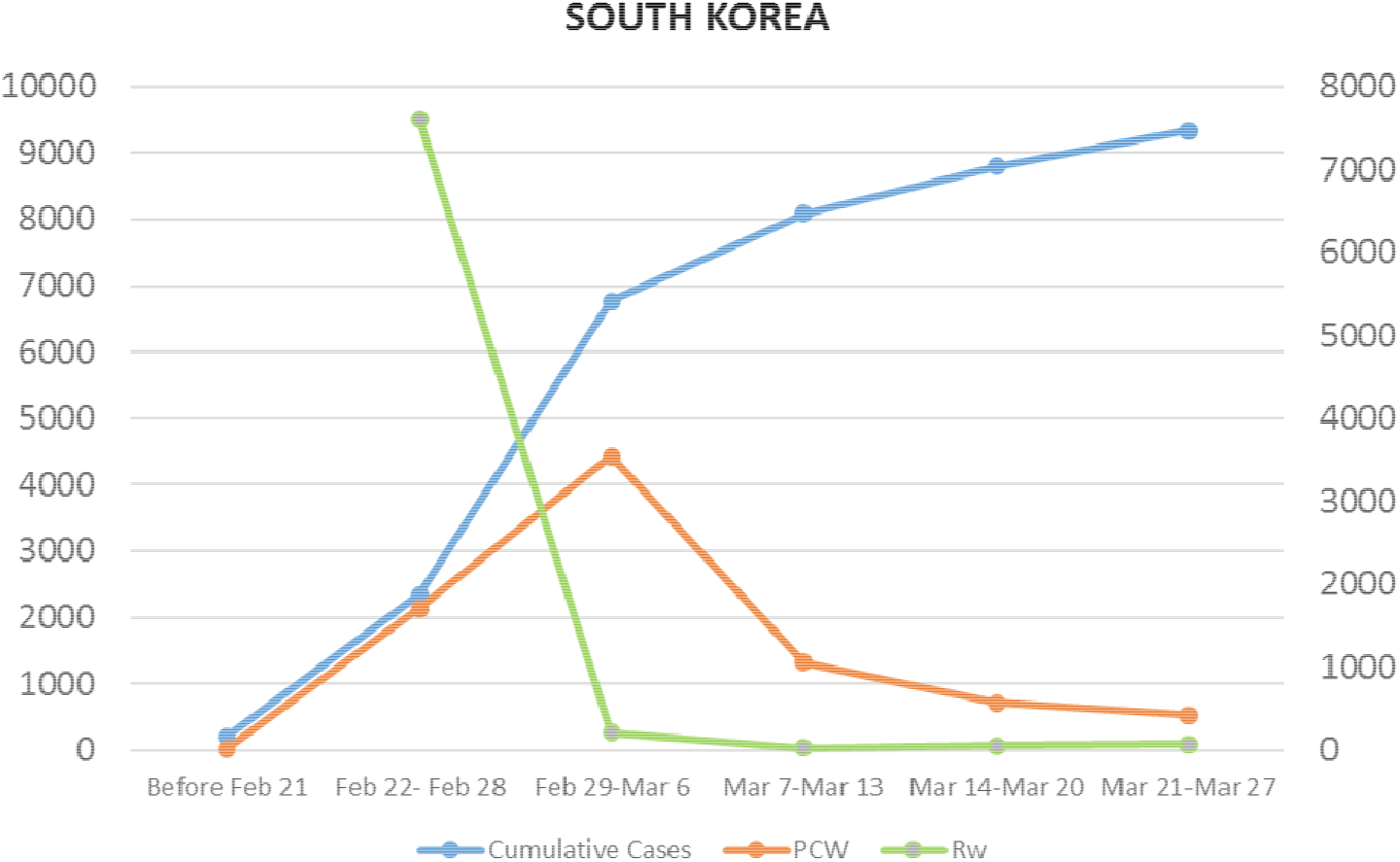
Graphical representation of positive cases for South Korea. (The secondary Y axis represents unit for)

### India

India reported its first case on January 30. It has passed Phase 1, which was from Feb 29- Mar 6. Now it’s in Phase 2, with a fluctuating value. It is predicted to be in Phase 2 for next two weeks. Positive cases reported have history of foreign travel or have come in contact with infected person. Community spread has not yet been reported from India. Coming weeks are crucial for containing COVID-19 pandemic. It is predicted to cross cumulative cases of 3,000 within Apr 3.

**Figure 10.**
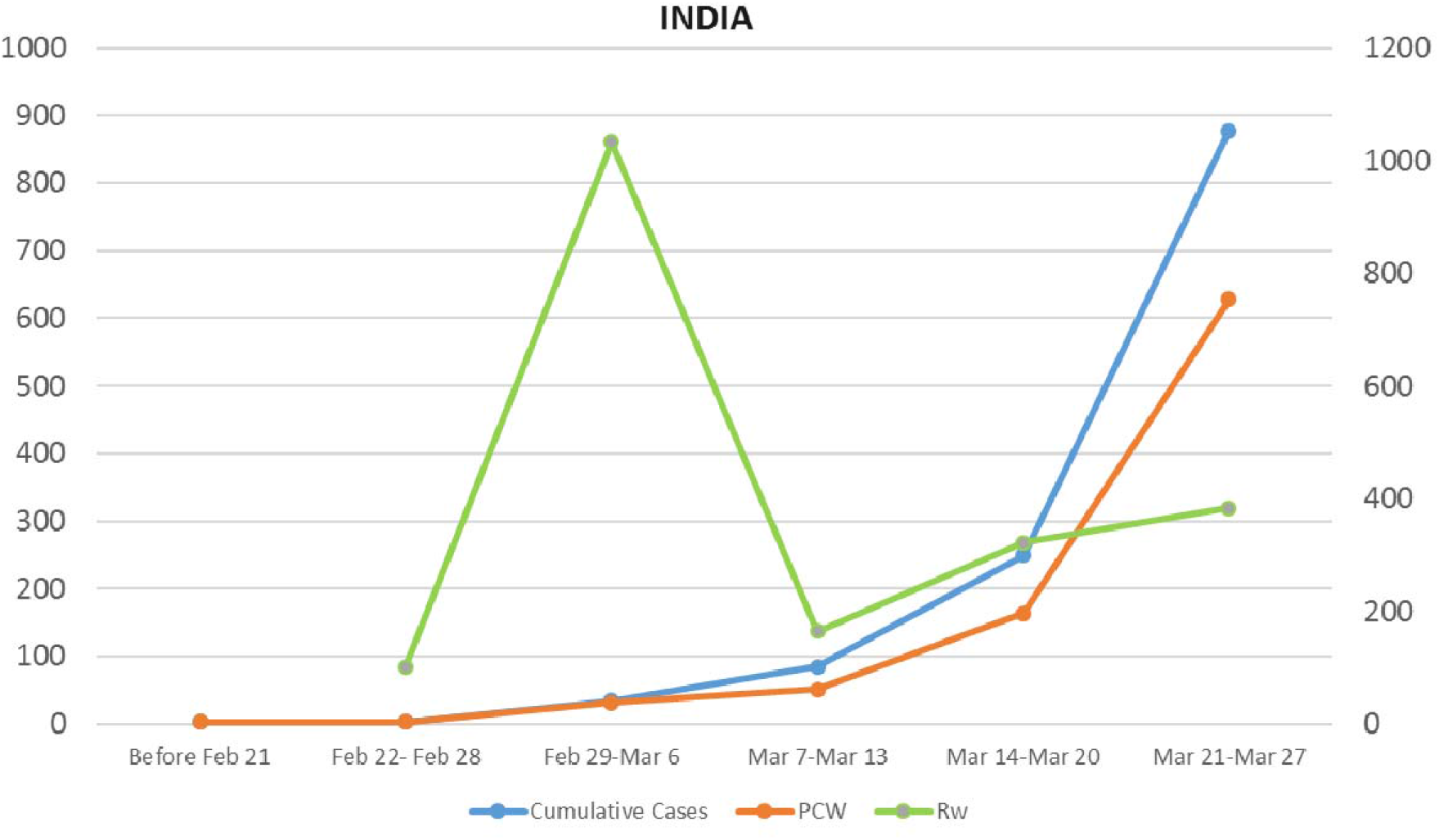
Graphical representation of positive cases for India. (The secondary Y axis represents unit for)

For the current COVID-19 pandemic, initial large-scale testing would help in identifying primary infected persons which would further help a country to transit from Phase 2 to Phase 3 rapidly, thus controlling the disease by identifying large section of infected people. Similarly, [16][17] suggested the importance of screening, surveillance and control efforts, particularly at airports and other travel hubs, in order to prevent further international spread of 2019-nCoV.

South Korea is a best example where for Phase 1 was highest 7617.86 among the countries under study, which was due to large scale testing and identification of positive case (7,290 person per million population as of Mar 27) [20], thus leading to quick transition from Phase 2 to Phase 3. Rapid, low-cost and large-scale testing methods for COVID-19 have been summarized by [17, 19] which can be used to mitigate such pandemic outbreak.

Italy, Iran, Spain and Germany are showing decreasing *R*_*w*_ and are expected to reach Phase 3 within next couple of weeks. Germany and Spain are now at the same *R*_*w*_ value, but Spain has higher number of positive cases than Germany. Italy, Iran and Germany had almost comparable number of positive cases 20,18 and 16 respectively as of Feb 21, but are now at cumulated positive cases of 86498, 32332 and 42288 respectively.

As of for US and India which are either fluctuating in Phase 2 or having slow decay in *R*_*w*_ value, should take immediate effective measures such as large-scale testing and self-isolation in order to reach Phase 3 in shortest period possible to reduce further exponential growth of positive cases.

### Limitations of Current method

The epidemic should show normal distribution. In the present method a week is considered to be from Saturday to Friday, changing this pattern to other forms have considerable effect on results. It is difficult to predict the magnitude of *PCW* but rather gives a fair direction of the Epidemic and can evaluate efficiency of control measures undertaken to combat the disease. This method is suitable for larger territories or regions with community spread of virus.

## Conclusion

The current method helps to understand the current state of Epidemic and can give fair prediction of Epidemic for upcoming weeks. Rapid and large-scale testing helps in controlling the COVID-19 outbreak. Initial screening and quarantining people with history of foreign travel helps in further to prevent spread of virus into community. As no vaccine has been developed up till now and control measures are the best option. Quick transition from Phase 2 to Phase 3 is crucial in controlling the COVID-19 pandemic.

## Data Availability

All the relevant data included in the manuscript.

## Acknowledgements

Author is immensely grateful to all resource persons for their useful advice.

## Funding

No funding agencies

## Conflict of Interest

None

## Supplementary Materials

**Supplementary Table 1.** Weekly *R*_*w*_ value from Feb. 22 to Mar. 27 for each Country.

|  | Italy | USA | Spain | Germany | Iran | France | South Korea | India |
| --- | --- | --- | --- | --- | --- | --- | --- | --- |
| **Feb 22 to Feb 28** | **4345.00** | 35.71 | 850.00 | 362.50 | **2055.56** | 375.00 | **7617.86** | 100.00 |
| **Feb 29 to Mar 6** | 431.19 | **4660.00** | 1079.41 | **793.10** | 1178.11 | **1235.56** | 207.69 | **1033.33** |
| **Mar 7 to Mar 13** | 347.58 | 820.17 | **1316.35** | 549.57 | 151.80 | 548.20 | 29.77 | 164.52 |
| **Mar 14 to Mar 20** | 225.44 | 799.32 | 338.21 | 430.97 | 125.13 | 293.668 | 54.0561 | 321.57 |
| **Mar 21 to Mar 27** | 134.45 | 542.42 | 260.04 | 260.04 | 153.24 | 227.37 | 74.75 | 382.93 |

The bold value indicates Phase 1 for respective Country.

## Notes

### Competing Interest Statement

The authors have declared no competing interest.

### Funding Statement

No external funding agency.

